# MRI-based classification of neuropsychiatric systemic lupus erythematosus patients with self-supervised contrastive learning

**DOI:** 10.1101/2021.04.16.21255634

**Authors:** Francesca Inglese, Minseon Kim, Gerda M. Steup-Beekman, Tom W.J. Huizinga, Mark van Buchem, Jeroen de Bresser, Daeshik Kim, Itamar Ronen

## Abstract

**Introduction/Purpose:** Systemic lupus erythematosus (SLE) is a chronic auto-immune disease with a broad spectrum of clinical presentations, including heterogeneous and uncommon neuropsychiatric (NP) syndromes. Accurate diagnosis of neuropsychiatric SLE (NPSLE) is challenging due to lack of clinically useful biomarkers. Despite structural brain abnormalities on MRI in NPSLE being a common finding, a robust link between structural abnormalities and NPSLE has not been established, thus their contribution to the distinction between NPSLE patients and patients in which the NP symptoms are not primarily attributed to SLE is limited. Self-supervised contrastive learning algorithms do not require labels, and have been shown to be useful in classification tasks in rare diseases with limited number of datasets. The aim of our study was to apply self-supervised contrastive learning on T_1_-weighted images acquired from a well-defined cohort of SLE patients to distinguish between SLE patients with NP symptoms due to the disease (NPSLE) or and SLE patients with similar symptoms due to other causes (non-NPSLE).

**Subjects and Methods:** 163 patients were included. We used 3T MRI T_1_-weighted images registered to the MNI152 template. The training set comprised 68 non-NPSLE and 34 NPSLE patients. During the training procedure, we applied random geometric transformations (cropping, left-right flipping and rotations) between iterations to enrich our data sets. Our ML pipeline consisted of convolutional base encoder and linear projector. To test the classification task, the projector was removed and one linear layer was measured. We trained the encoder and projector with the Normalized Temperature-scaled Cross Entropy Loss (NT-xent) loss function. We performed a Monte Carlo validation that consisted of 6 repeated random sub-samplings each using a random selection of a small group of samples from each group.

**Results:** In the 6 trials described above, between 79% and 83% of the patients were correctly classified as NPSLE or non-NPSLE. For a qualitative evaluation of spatial distribution of the common features found in the NPSLE population, Gradient-weighted Class Activation Maps (Grad-CAM) were examined voxel-wise. Thresholded Grad-CAM maps show areas of common features identified for the NPSLE cohort, with no such communality found for the non-NPSLE group.

**Discussion/conclusion:** The self-supervised contrastive learning model was effective in capturing diagnostic brain MRI features from a limited but well-defined cohort of SLE patients with NP symptoms. The interpretation of the Grad-CAM results is not straightforward, but points to involvement of the lateral and third ventricles, periventricular white matter and basal cisterns. We believe that the common features found in the NPSLE population in this study indicate a combination of tissue loss, local atrophy and to some extent that of periventricular white matter lesions, which are commonly found in NPSLE patients and appear hypointense on T_1_-weighted images.

## 1 Introduction

Systemic lupus erythematosus (SLE) is a female-predominant auto-immune disease with a broad spectrum of clinical presentations and multi-organ involvement. SLE is characterized by the production and deposition of several autoantibodies [1]. The involvement of the central nervous system (CNS) in SLE leads to a series of non-specific neuropsychiatric (NP) manifestations in between 12-95% of SLE patients [2]. These NP symptoms widely range in terms of severity and prognostic implications [3]. NP events in SLE can be directly associated with the disease (NPSLE) or can be explained by another aetiology (non-NPSLE). NP symptoms are associated with an increased mortality and reduced quality of life within the SLE population [4]. TThe diagnosis of NPSLE is also difficult due to the heterogeneous nature of NP syndromes, which include headache, seizures, anxiety and psychosis [5], and the large variation in the attribution of the NP symptoms across studies and institutions highlights the difficulty in unequivocally diagnose NPSLE. In clinical practice, it is important to correctly classify NP events, since the therapeutic approach is defined based on this classification. A study performed in our center reported that about 15% of NP events attributed to SLE (NPSLE) during the first patient evaluation were reclassified after reassessment as non-NPSLE [6]. This discrepancy highlights the pressing need for biomarkers that will contribute to more reliably distinguish between NPSLE and non-NPSLE early in the diagnostic process.

Two different underlying mechanisms are thought to play a role in the pathophysiology of NPSLE. One is the inflammatory mechanism, where the blood-brain barrier (BBB) or the blood-cerebrospinal fluid (BCSF) barrier [7] is compromised due to presence of pro-inflammatory factors. Subsequently, auto-antibodies can enter the brain and trigger an inflammatory process that results in focal or diffuse tissue damage. The second proposed mechanism is the thrombotic or ischemic mechanism, where vascular injury and occlusion are present [8]. Due to the lack of a gold standard for the diagnosis, the best strategy so far for diagnosing NPSLE remains a multidisciplinary expert consensus after standardized evaluation of complaints and a complete battery of tests, including brain magnetic resonance imaging (MRI) [6]. Despite conventional brain MRI being the method of choice for clinical evaluation of SLE patients experiencing NP events, morphological changes and brain lesions observed in these patients do not clearly correlate with the clinical symptoms and disease outcome, underscoring the clinical-radiological paradox encountered with many NPSLE patients, defined by the presence of lesions in the absence of symptoms of NPSLE or viceversa [9].

Currently, MRI features can only contribute in a limited way in the diagnostic process, mostly in the way of exclusion. Several studies have shown that patients with SLE have more white matter hyperintensities (WMH) and more atrophy and infarcts compared to controls [10, 11, 12, 13, 14]. These findings per se, albeit indicative of robust presence of structural abnormalities in NPSLE, are not useful yet for the diagnostic process. Mostly – basic metrics such as global atrophy and lesion count/load do not lead to a specific diagnosis. It is therefore imperative to further explore neuroimaging biomarkers that can help clinicians to differentiate between NPSLE and non-NPSLE patients, and further down the line, also help in the stratification of NPSLE patients based on their clinical phenotype.

Deep learning has been shown to be useful in diagnostic tasks related to clinical neuroimaging data both in diseases with overt brain damage such as stroke, as well as in diseases in which brain alterations are not directly detectable in standard radiological observation [15]. Deep learning models can extract significant features that are relevant in clinical diagnosis and can distinguish between patient populations even when the brain alterations are not visibly overt [16]. Classification tasks, however, require a large number of data sets, as well as trained clinicians to generate labels to aid the categorization process. Dementia [17, 18, 19, 20] and psychiatric disorders [21, 22] have been natural targets for using deep neural networks as imaging data for these diseases are widely available. NPSLE, on the other hand, is a sub-category of SLE, which in itself is categorized as an orphan/rare disease (prevalence of 1-5 in 10,000, source: www.orpha.net) and thus the amount of data available is limited. This makes a supervised ML approach impractical for studying brain abnormalities in NPSLE in a single-center study.

Recently, self-supervised learning approaches, which train the model on unlabeled data by providing self-generated labels from the data themselves, have become popular in image classification. In non-medical applications, self-supervised learning approaches were applied in the prediction of the rotation angles of objects [23], colorization of gray-scale images [24] and solving randomly generated Jigsaw puzzles [25]. Instance-level identity preservation with contrastive learning has shown to be effective in learning rich representations for classification [26, 27, 28]. In this context, self-supervised learning approaches are more suitable for dealing with limited data sets, such as the one presented in this work. Self-supervised learning in biomedical imaging has been implemented in several instances, among which screening of 2-dimensional chest x-ray images [29], in the evaluation of cardiac time-series data [30], in tissue segmentation of brain lesions [31], in segmentation of the renal dynamic contrast-enhanced MRI [32], in robust and accelerated reconstruction of quantitative and B_0_-inhomogeneity-corrected R _*\**2_ maps from multi-gradient recalled echo MRI data [33] and in quality enhancement of compressed sensing MRI of vessel wall [34].

In this study, we hypothesized that a self-supervised learning approach would be effective for the classification tasks in our limited patient population, in particular in the distinction between two important diagnostically different SLE patient groups: NPSLE and non-NPSLE patients. To test this hypothesis, we applied a self-supervised method to 3D structural MRI data with the aim of distinguishing and classifying such data for NPSLE and non-NPSLE patients.

## 2 Methods

### 2.1 Patient population

The Leiden University Medical Center (LUMC) is the national referral center in the Netherlands for SLE patients with NP complaints. SLE Patients are referred to the outpatient clinic if they present with NP manifestations. In this retrospective study we included 216 patients with SLE recorded between May 2007 and April 2015. Of these, 28 patients were excluded because of undefined diagnosis, 3 patients were excluded because of motion artefacts in the MRI scan, 20 patients were excluded because of brain infarcts over 1.5 cm and 2 patients were excluded due to the presence of other diseases (one for a brain tumor and one for a large arachnoid cyst). This resulted in a total of 163 patients included in this study. The medical ethics committee of Leiden-The Hague-Delft approved of the study and all included patients signed for informed consent.

All patients came to the clinic for a full one-day visit and underwent an identical standardized assessment that included a brain MRI scan [35] and a combination of multidisciplinary medical assessments and extensive complementary tests, necessary for deciding whether the NP-events are attributed to SLE [8]. Attribution of NP symptoms to SLE was established during a multidisciplinary consensus meeting. This diagnostic process is described in detail previously [35]. NP events were classified according to the 1999 ACR nomenclature for NPSLE [36].

During an intake interview, information about gender, age, and SLE disease duration was provided by the patients and verified by their medical records. During the evaluation, SLE activity and damage indexes were scored for each patient: the SLE disease activity was defined using the Systemic Lupus Erythematosus Disease Activity Index 2000 (SLEDAI-2K) [37]; SLE irreversible damagewas determined through the Systemic Lupus International Collaborating Clinics/American College of Rheumatology damage index (SDI) [38].

### 2.2 MRI protocol

All patients were scanned, according to a standardized scanning protocol, on a Philips Achieva 3T MRI scanner (Philips Healthcare, Best, The Netherlands) with a body transmit RF coils and an 8-Channel head coil array. The sequence used for this project was a 3D T_1_-weighted gradient echo scan (voxel size= 1.17 *×* 1.17 *×* 1.2`mm`^3^; TR/TE= 9.8*/*4.6`ms`).

### 2.3 MRI preprocessing

All the T1-weighted images were registered to a standard brain template, the Montreal Neurological Institute standard template (MNI152), using FNIRT (FMRIB’s non-linear image registration tool) [39, 40], using affine registration with 12 degrees of freedom.

### 2.4 Machine learning pipeline architecture

We followed the self-supervised framework introduced by Chen et al. [26], where an encoder network *f*_*θ*_ was used to project the image into a feature space, followed by two-layer multi-perceptron (MLP) *p*_*π*_ projector that projected the features into latent vector *z*. In our work we modified this approach to address the fact that 3D MRI data requires more classification parameters than 2D natural images. Therefore, we designed the encoder network *f*_*θ*_ using three convolutional layers with batch normalization and a max pooling layer. To test the representation feature, we changed the projector layer into a linear layer which had the same output size as the number of classes, which in our case equals 2 – NPSLE and non-NPSLE. Subsequently, we fine-tuned the linear layer with a training set. Finally, we tested the accuracy of the trained encoder and linear layer. Figure 1 shows the architecture described above.

**Figure 1:**
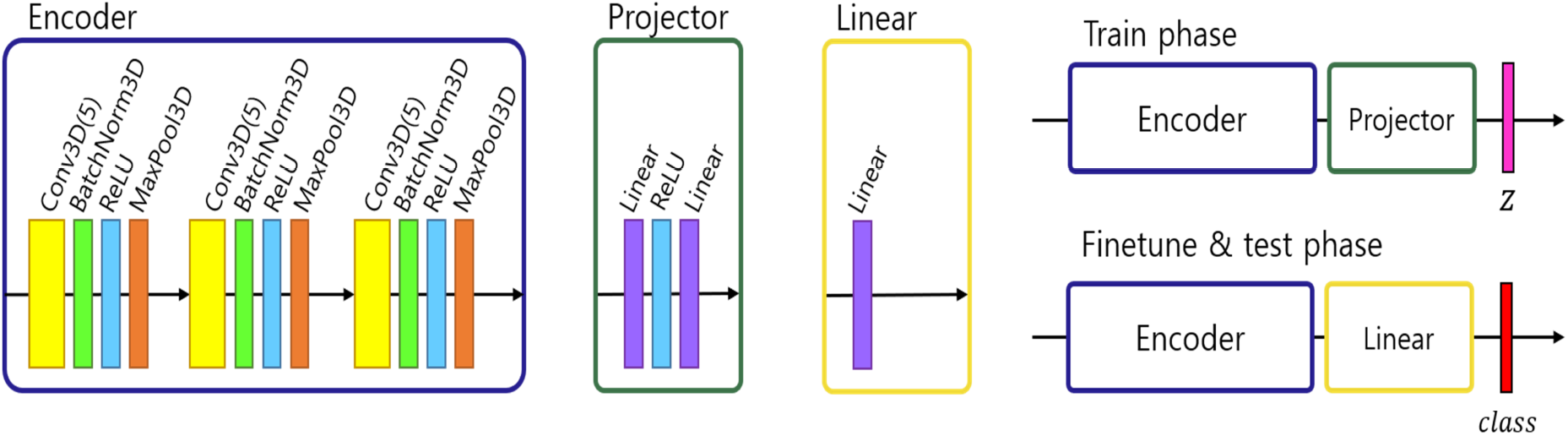
Architecture of ML pipeline: encoder, projector and linear module. In the training phase, an encoder and projector were used to project the images into representation space with a latent vector z. In the fine tuning and test phase, the projector was changed with a linear module and predict the class.

### 2.5 Preprocessing

In 2D natural images, Chen et al. [26] used stochastic data augmentation *t*, randomly selected from the family of augmentation T, including random cropping, random color distortion, and random flip. In our 3D MRI data, we used stochastic data augmentation, by performing random selection from the set of augmentations we applied to our data. These included: random cropping, random flipping along the *z* axis and random in-plane rotation. The random crop was applied up to 15 voxels along the three axes. For the random flip, only left-right flips were applied based on the foot-head axis (coronal plane). For rotation, random rotation of angles up to 45 degrees was applied in the left-right, anterior-posterior and foot-head directions. To test the robustness of our method with respect to different data augmentation strategies, we tried three different augmentation settings: one only with crop, one with crop, flip and rotation. The geometric transformations are depicted in Figure 2.

**Figure 2:**
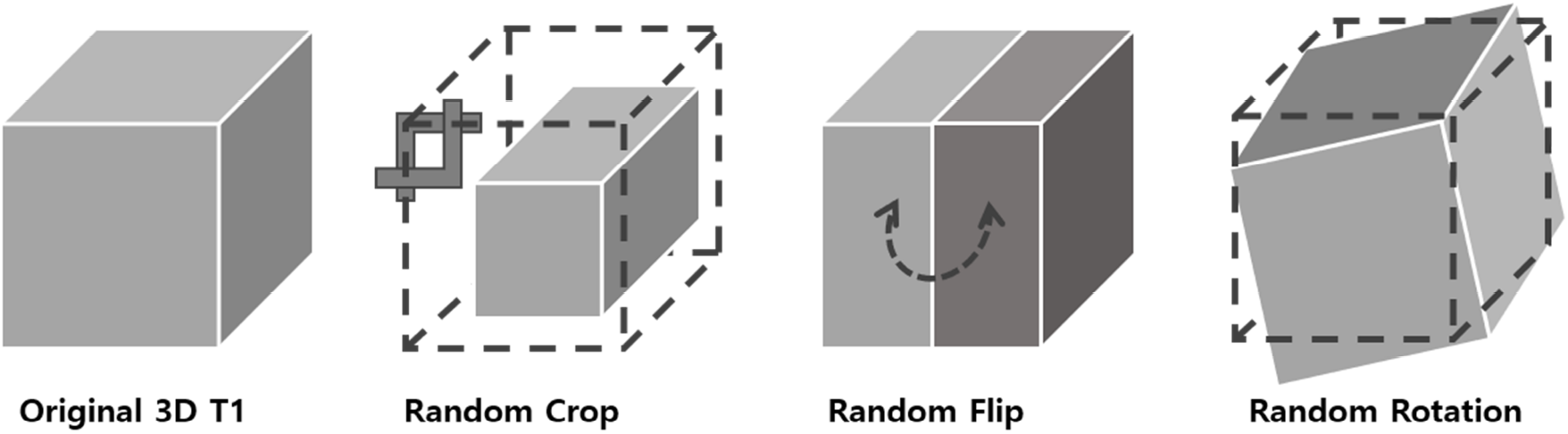
3D data transformation. Three types of geometrical transformations were applied to the 3D MRI data augment the data set: cropping of the image, left-right flips, and in-plane rotations.

### 2.6 Contrastive loss

The contrastive loss function *ℒ*_con_ is defined as follows,

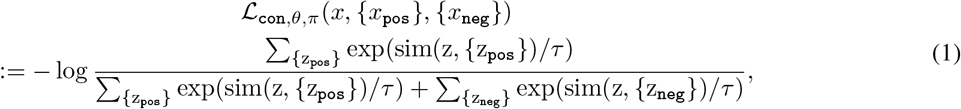

where *z*, {*z*_*pos*_}, and {*z*_*neg*_}are corresponding 128 dimensional representation vectors (*z*) of *x* obtained by the encoder and projector *z* = *p*_*π*_ (*f*_*θ*_ (*x*)). The sim(*u, v*) = *u*^*T*^ *v*/ ‖*u*‖ ‖*v*‖ denotes cosine similarity between two vectors and *τ* is a temperature parameter [26]. We trained the encoder and projector with the contrastive loss function, *NT-xent* which maximizes the similarity between each transformed sample.

### 2.7 Patients selection for validation of the ML pipeline

To determine the accuracy of our study, six trials were performed. In each trial, the training set consisted of 68 non-NPSLE and 34 NPSLE patients randomly chosen from within the total patient population (163 subjects). In order to have an equal number of NPSLE and non-NPSLE patients for the training procedure, we used the images of NPSLE twice in every epoch. Therefore, a total of 136 images were used to train the model. In each trial, the test set consisted of 9 non-NPSLE patients and 9 NPSLE patients randomly chosen within the total patient’s population (163 subjects) excluding the training set. The demographic and clinical characteristics of the patients selected in each trial are shown in Tables 1.

**Table 1:**
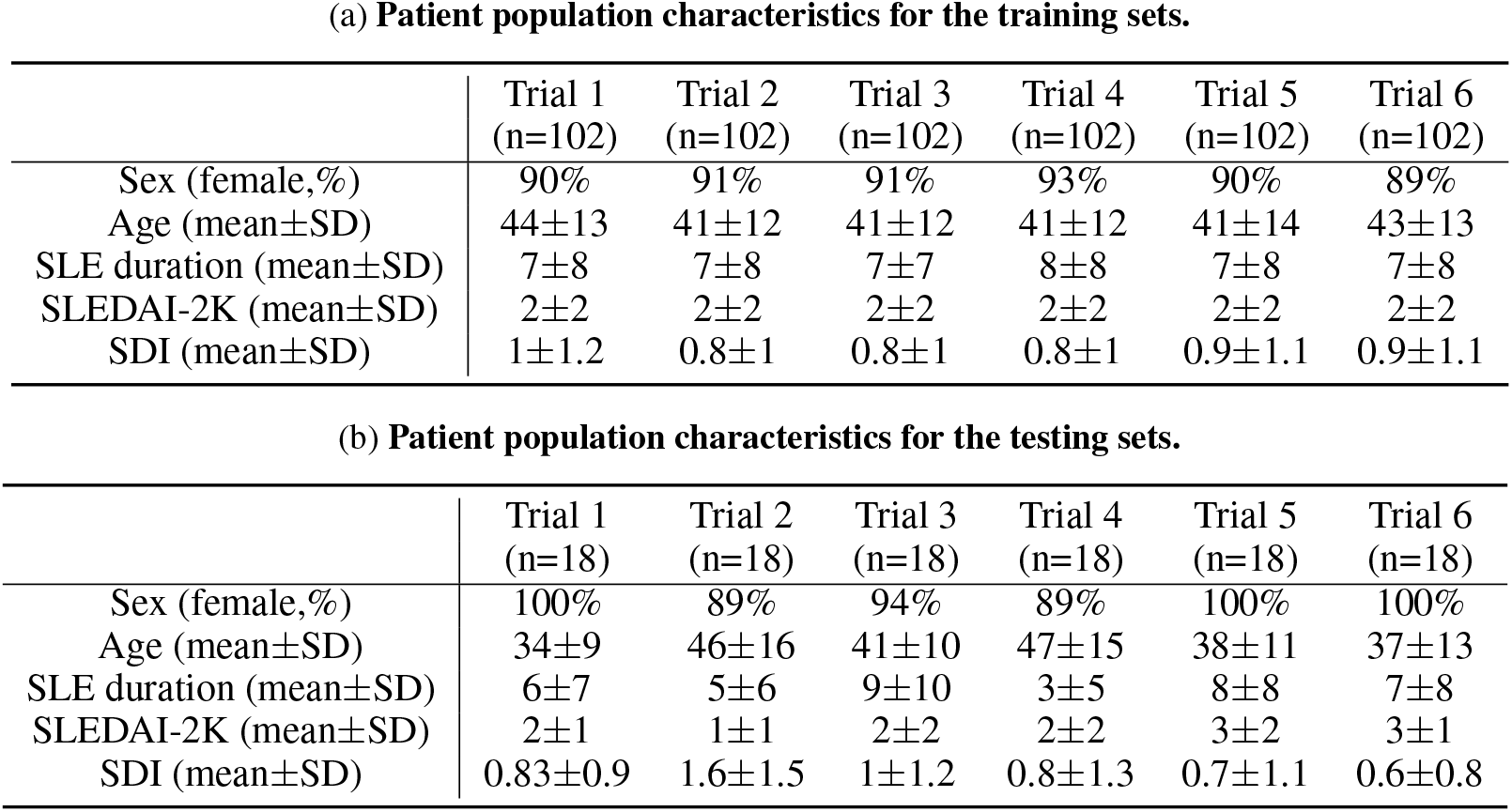
Patient population characteristics. Sex, age and SLE clinical variables are described for each trial. SLEDAI= systemic Lupus Erythematosus Disease Activity Index 2000. SDI= Systemic Lupus International Collaborating Clinics/American College of Rheumatology Damage Index.

|  | Trial 1<br>(n=102) | Trial 2<br>(n=102) | Trial 3<br>(n=102) | Trial 4<br>(n=102) | Trial 5<br>(n=102) | Trial 6<br>(n=102) |
| --- | --- | --- | --- | --- | --- | --- |
| Sex (female,%) | 90% | 91% | 91% | 93% | 90% | 89% |
| Age (mean±SD) | 44±13 | 41±12 | 41±12 | 41±12 | 41±14 | 43±13 |
| SLE duration (mean±SD) | 7±8 | 7±8 | 7±7 | 8±8 | 7±8 | 7±8 |
| SLEDAI-2K (mean±SD) | 2±2 | 2±2 | 2±2 | 2±2 | 2±2 | 2±2 |
| SDI (mean±SD) | 1±1.2 | 0.8±1 | 0.8±1 | 0.8±1 | 0.9±1.1 | 0.9±1.1 |

(b) Patient population characteristics for the testing sets.
|  | Trial 1<br>(n=18) | Trial 2<br>(n=18) | Trial 3<br>(n=18) | Trial 4<br>(n=18) | Trial 5<br>(n=18) | Trial 6<br>(n=18) |
| --- | --- | --- | --- | --- | --- | --- |
| Sex (female,%) | 100% | 89% | 94% | 89% | 100% | 100% |
| Age (mean±SD) | 34±9 | 46±16 | 41±10 | 47±15 | 38±11 | 37±13 |
| SLE duration (mean±SD) | 6±7 | 5±6 | 9±10 | 3±5 | 8±8 | 7±8 |
| SLEDAI-2K (mean±SD) | 2±1 | 1±1 | 2±2 | 2±2 | 3±2 | 3±1 |
| SDI (mean±SD) | 0.83±0.9 | 1.6±1.5 | 1±1.2 | 0.8±1.3 | 0.7±1.1 | 0.6±0.8 |

### 2.8 Training details

We used three convolutional layers as the base encoder network and 2-layer multi-layer perceptron with 128 embedding dimensions as the projection head. All models were trained by minimizing the final contrastive loss with a temperature of *τ* = 0.5. For the rest, we followed similar optimization steps as in SimCLR [26]. We trained with 1,000 epochs under the stochastic gradient descent (SGD) base Layer-wise Adaptive Rate Scaling (LARS) optimizer [41], a cosine annealing learning rate and a gradual warmup scheduler [26]. We used a weight decay of 1*e* − 6 and momentum of 0.9. We used linear warm-up for the first 10 epochs until a learning rate of 1.0 was achieved and decay with cosine decay schedule. We used a batch size of 16. Furthermore, we used global batch normalization, which shared the parameters over the multiple GPUs. We trained the encoder with contrastive loss. Then, we fine-tuned the encoder along a linear layer to optimize the cross-entropy loss with learning rate 0.001 during 35 epochs.

### 2.9 Gradient class activation mapping

There are several methods for interpreting predictions in deep learning [42, 43, 44, 45, 46, 47]. We chose to use gradient class activation mapping (grad-CAM) [42]. Grad-CAM uses gradient information flowing into the last convolutional layer of the model to assign importance values to each parameter for the prediction of the model. We extended the grad-CAM to deal with 3D convolutional neural networks. Since we are interested in a classification for a specific disease, and images are all aligned to the same space (MNI 152), it was possible to average the grad-CAM maps of test samples to obtain a qualitative measure of the brain regions where significant features were found after averaging across subjects in each trial. For this purpose, for each trial, the grad-cam maps of the samples which were correctly classified in the same class were averaged. Thresholded averaged grad-CAM maps were then overlaid on the MNI template for display.

### 2.10 Statistics

For evaluation of the performance of the algorithm at each trial, we used classification *accuracy, precision* and *recall* as quantitative measures for the performance of the algorithm. An image is considered correctly classified when the model classifies it into the correct class: NPSLE or non-NPSLE. Accuracy was defined as the ratio between the total number of correctly classified samples and the total number of test sets. The coefficient of variation for the three augmentation strategies was also calculated by taking the ratio of the mean accuracy and the standard deviation. Precision was defined as the fraction of correctly predicted NPSLE samples out of the total test samples (NPSLE + non-NPSLE). The recall, defined as the fraction of relevant items selected from the interest class, as calculated as the fraction of correctly predicted NPSLE samples out of the total number of NPSLE samples.

To assess differences in mean accuracy, precision and recall among the three different geometric transformation approaches we used here, repeated-measures analysis of variance (ANOVA) was performed using the Statistical Package for the Social Sciences (SPSS) version 25 (IBM corporation, Armonk, NY, USA).

## 3 Results

### 3.1 Classification performance

Table 2a shows the individual and mean accuracy of the classification results for the six trials, defined as the percent of correctly classified NPSLE/non-NPSLE patients out of the total tests within a trial. Results are given for three different strategies for data augmentation that included one (random crop), two (random crop + flip) and three (crop + flip + rotate) transformations. The accuracy(SD) of the six trials in each augmentation strategy was 83.35% (14.05%), 83.33% (16.10%) and 79.63% (11.47%). No significant differences were found when comparing the accuracy of the classification across the three data augmentation options. The coefficient of variation for the three augmentation strategies ranged between 0.14 and 0.19.

**Table 2:** Results of accuracy, precision and recall.

| Random data augmentation Set | T | Trial 1 | Trial 2 | Trial 3 | Trial 4 | Trial 5 | Trial 6 | Percentage (SD) |
| --- | --- | --- | --- | --- | --- | --- | --- | --- |
| Crop | $\{T_1\} \in T$ | 61.11<br>(6,5) | 77.78<br>(8,6) | 77.78<br>(5,9) | 100.0<br>(9,9) | 88.89<br>(7,9) | 94.44<br>(9,8) | 83.35<br>(14.05) |
| Crop, Flip | $\{T_1, T_2\} \in T$ | 61.11<br>(7,4) | 66.67<br>(6,6) | 94.44<br>(8,9) | 83.33<br>(7,8) | 94.44<br>(8,9) | 100.00<br>(9,9) | 83.33<br>(16.10) |
| Crop, Flip, Rotation | $\{T_1, T_2, T_3\} \in T$ | 66.67<br>(6,6) | 72.22<br>(8,5) | 77.78<br>(6,8) | 100.0<br>(9,9) | 77.78<br>(8,6) | 79.63<br>(7,8) | 79.63<br>(11.47) |

| Random data augmentation Set | T | Trial 1 | Trial 2 | Trial 3 | Trial 4 | Trial 5 | Trial 6 | Percentage (SD) |
| --- | --- | --- | --- | --- | --- | --- | --- | --- |
| Crop | $\{T_1\} \in T$ | 45.5/<br>55.6 | 42.9/<br>66.7 | 64.3/<br>100.0 | 50.0/<br>100.0 | 56.3/<br>100.0 | 44.4/<br>88.9 | 50.6 (8.0)/<br>85.2 (19.0) |
| Crop, Flip | $\{T_1, T_2\} \in T$ | 36.4/<br>44.4 | 50.0/<br>66.7 | 52.9/<br>100.0 | 53.3/<br>88.9 | 52.9/<br>100.0 | 50.0/<br>100.0 | 49.3 (6.0)/<br>83.3 (23.0) |
| Crop, Flip, Rotation | $\{T_1, T_2, T_3\} \in T$ | 50.0/<br>66.7 | 38.5/<br>55.6 | 57.1/<br>88.9 | 50.0/<br>100.0 | 42.9/<br>66.7 | 52.9/<br>100.0 | 48.6 (7)/<br>79.7 (19) |

Table 2b shows the individual and mean precision and recall of the 6 classification trials. Precision is defined as percent of correctly predicted NPSLE cases out of the total number of tests within a trial, and recall is defined as the fraction of correctly predicted NPSLE samples out of the total number of NPSLE test samples within a trial. The same three sets of geometric transformation for data augmentation were used also here. The precision (SD) across trials was 50.6% (8%), 49.3% (6%), and 48.6% (7%) for trials using only crop, crop + flip, and crop + flip + rotation transformation, respectively. Recall (SD) were 85.2% (19%), 83.3% (23%), and 79.7% (19%) for trials using only crop, crop + flip, and crop + flip + rotation transformation, respectively.

No significant differences were found in accuracy (p=0.531), precision (p=0.845) and recall (p=0.686) among trials using the 3 different geometric transformations.

### 3.2 Common features in NPSLE - Grad-CAM results

Figures 3a through 3c show the thresholded averaged grad-CAM maps for the 6 trials. Threshold was set at three different levels: 0.75, 0.85 and 0.95. Each of the three figures shows the results following data augmentation with crop only (3a), crop + flip (3b) and crop + flip + rotate (3c). Common features that show up on the grad-CAM maps were found only in the NPSLE cohort. The areas generated by the grad-CAM maps with threshold set at 0.75 are too generic to report on specific brain regions, while the higher thresholds of 0.85 and 0.95 reveal more parcellated maps indicating local involvement. Brain regions included in the grad-CAM maps with threshold above 0.85 are lateral ventricles and periventricular white matter, as well as third ventricle and basal cisterns (Figure 4). There were no specific regions that contributed to the model’s prediction of non-NPSLE.

**Figure 3:**
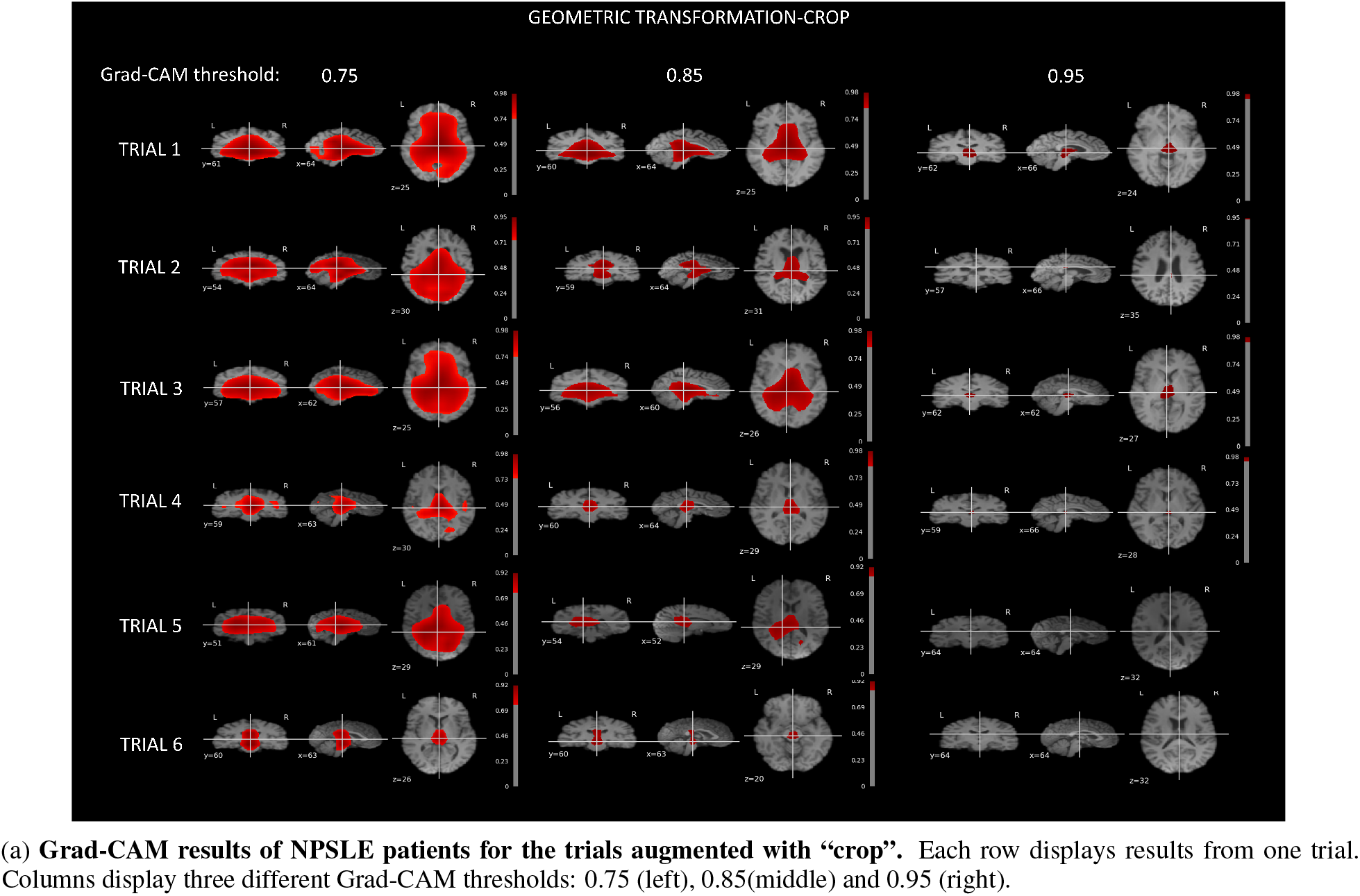
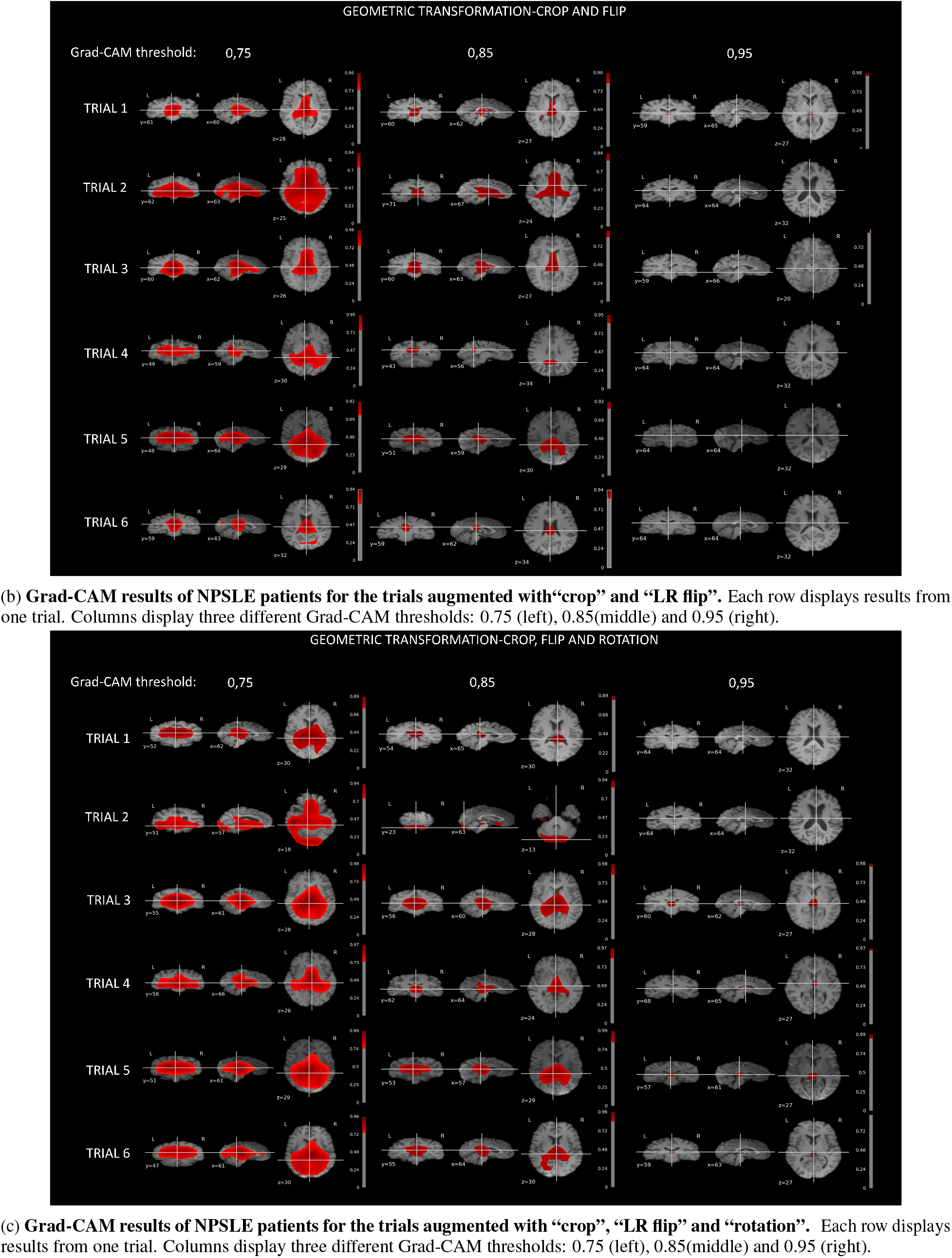
Grad-CAM results with different data augmentation set.

**Figure 4:**
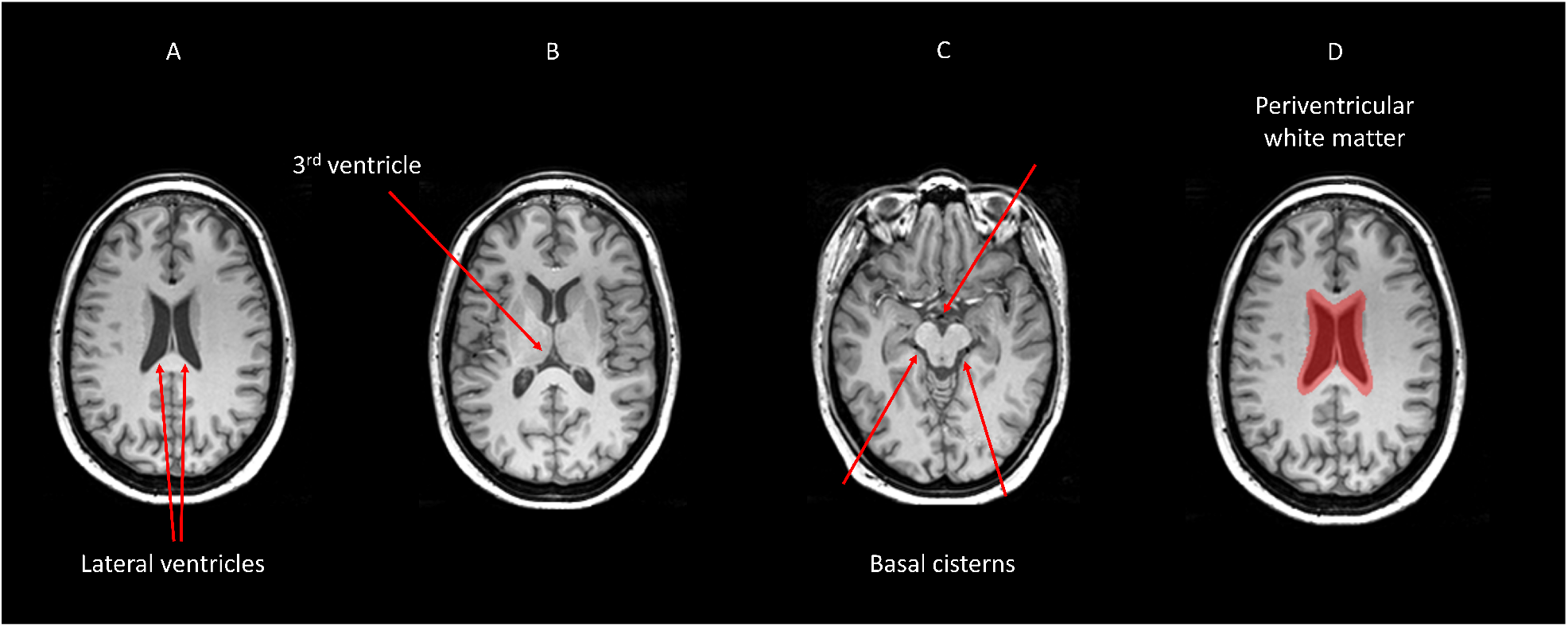
Brain areas that correspond with regions highlighted in the grad-CAM maps with threshold above 0.85.

## 4 Discussions

In this study, we designed a self-supervised machine-learning pipeline for classification of T1-weighted MRI images, aimed at distinguishing between images of NPSLE patients and those of non-NPSLE patients. The accuracy of the classification algorithm, based on 6 repeated trials, was significantly above random choice, and practically independent of the augmentation strategy. The mean classification accuracy into the two classes, NPSLE and non-NPSLE, across the three augmentation strategies ranged between 79% and 83%. Within-augmentation variability across trials was well contained: the coefficient of variation for the three augmentation strategies ranged between 0.14 and 0.19, indicating a good repeatability of the accuracy of the classification, despite the significant heterogeneity of clinical measures and the symptoms in the NPSLE population.

The overall precision of our model, representing the fraction of correctly predicted NPSLE out of the total number of NPSLE predictions, was lower, at about 51.8%. This indicates a relatively high rate of false negatives (non-NPSLE subjects identified as NPSLE). This may indicate that the structural brain changes characteristic of NPSLE and were picked up by the classification algorithm can also be found in non-NPSLE patients. MRI abnormalities, such as lesions, local atrophy and other diffuse abnormalities have been found in non-NPSLE and as well as in SLE patients without NP when compared with healthy controls [10, 11, 12, 13, 14]. It is thus not surprising that some of the features picked up in the training of the algorithm were erroneously attributed to non-NPSLE patients. It remains to be seen whether with increased population size (via, for example, a multicenter effort), or additional MRI modalities that reflect better other aspects of structural changes in the disease (for example, FLAIR T2 images that report on white matter hyperintensities) will contribute to the precision of the classification and limit the false-positives. Conversely, the recall, defined as the fraction of total relevant results (correctly predicted NPSLE patients) out of the NPSLE group, averaged at about 83% indicating a relatively low rate of false negatives, i.e. NPSLE patients that were not classified as such. This corroborates the meaningfulness of the features found by the ML algorithm and provides support to their link to brain changes in NPSLE population.

We studied the relationship between brain alterations in NPSLE and the common features identified by the classification algorithm with grad-CAM, a commonly used visualization tool that provides a coarse localization map highlighting important regions in the image for the classification task. As of recent, Grad-CAM maps have been applied to medical imaging modalities, including a successful application aimed at grading gliomas based on MR images [48, 49]. While grad-CAM maps do not provide quantitative statistical information in the way that statistical parametric maps do, they do indicate communality in the features that led to the classification. In particular, basal cistern, lateral ventricles, third ventricle and periventricular white matter seemed to be features that discriminate NPSLE patients from non-NPSLE. Studies on structural MRI pre [50] and post-mortem [51] of brains of NPSLE patients showed small focal lesions and white matter hyperintensities concentrating on periventricular white matter, as well as ventricular dilation. Higher occurrence of periventricular and deep WMH lesions was reported in SLE patients compared to controls but without stratification for NPSLE versus non-NPSLE patients [52]. Overall, the etiology of periventricular WMH in NSPLE is not well understood, not is it fully investigated in other diseases with prevalence of periventricular WMH. In older adults, periventricular WMH appear to be associated with impaired cognitive function, in particular with working memory, and are linked to disruption of long distance white matter connections. Some characterization of periventricular WMH in older adults was provided by diffusion tensor imaging and pathological observations and revealed that periventricular WMH are mostly characterized by gliosis and myelin loss [53]. Further investigation on the role of periventricular WMH in NPSLE patients is necessary to confirm their role and their importance in the disease process. There is no consistent reporting on direct involvement of the basal cisterns in SLE or NPSLE, barred few case reports [54, 55]. A plausible explanation for the involvement of the basal cisterns is the effect of atrophy, leading to increase in CSF volume in the basal cisterns as well as in the lateral ventricles.

There are several limitations and challenges associated with our study, and we hope to address some of them in our future efforts. Characterization of the overlapping features between the false negatives and “true” NPSLE subjects will certainly require special attention in future studies, with significantly larger populations of SLE and non-NPSLE patients, and with an inclusion of control groups of SLE patients without NP, as well as of healthy controls.

Despite the fact that there were common regions highlighted by Grad-CAM in the NPSLE group, it is premature to claim at this point that these regions are indeed clinically significant, and a broader investigation is required. In the current investigation we used only one MRI modality, namely T_1_-weighted images, a modality that is highly sensitive to volumetric structural changes, but less sensitive and less specific to lesions, infarcts, microbleeds and hemorrhages, which all result in local hypointensities. Thus, it is imperative to continue the investigation with a more multimodal approach, with additional modalities that will add more sensitivity and specificity to a variety of structural changes commonly found in SLE and NPSLE. From the algorithm perspective, self-supervised machine learning is identity preserving, and it is therefore possible to add a variety of MRI modalities to the process in the hope of significantly improving the classification performance.

In deep learning classification tasks, a large amount of data sets is essential. The number of samples (overall number of patient data sets) that were included in our study (163) was limited compared to typical numbers of samples used in classification tasks, typically in the tens of thousands of cases [29]. NPSLE is a rare and highly heterogeneous disease and it is therefore not a natural target for unsupervised machine learning based classification. The goal of this study was to establish a retrospective link between (known) diagnosis and structural brain differences between two classes of samples: NPSLE and non-NPSLE patients. Within the limits of a single-center study we benefitted from the maximum number of patients available in the Netherlands, as well as from the most comprehensive diagnostic process for NPSLE (being a national referral center for the disease). When attempting to use a 2 class-wise supervised learning approach, the model diverged, possibly due to the limited number of data sets and the inconspicuousness of the visual features. Based on the fact that self-supervised learning is known to perform well even with a small data sets [56, 57], we opted for self-supervised learning, in the hope that following the classification, common features for two independent classes will emerge, coinciding with the NPSLE and non-NPSLE patient groups. Eventually, the algorithm was only able to find common features within one class of patients (NPSLE), and no common features were found in the non-NPSLE group in our trials. We cannot claim as a certainty that there are no common features to the non-NPSLE group, but the positive result we obtained for the classification of the NPSLE group supports the notion that in our classification task it was more effective to allow the algorithm to learn the visual representation in a self-supervised manner using similarity across images, rather than providing a-priori class labels.

To conclude: we set the stage for classification of brain imaging data of NPSLE and non-NPSLE patients using deep neural networks, achieving relatively high average accuracy across repeated trials. We showed that self-supervised learning is capable of capturing common image features in one class of subjects (NPSLE), a task that is impossible to accomplish with supervised learning, that can capture relatively inconspicuous visual information by cross-entropy loss in the MRI images, and is advantageous when only a limited number of data sets is available. The method is modular and can accommodate additional imaging modalities to be included in the classification, and can be easily applied to other studies of rare diseases that suffer from similar limitations.

## Data Availability

Anonymized MRI data are available from Itamar Ronen subject to data transfer agreement.

## Acknowledgement

We thank Dr. Sahar Yousefi for her critical reading of the manuscript and her useful comments on the ML methodological aspects. We also thank Jihoon Tack for his help in implementing the deep learning model and useful comments on the ML development aspects.

